# Efficacy and Safety of Antibiotic Agents in Children with COVID-19: A Rapid Review

**DOI:** 10.1101/2020.04.13.20064402

**Authors:** Jianjian Wang, Yuyi Tang, Yanfang Ma, Qi Zhou, Weiguo Li, Muna Baskota, Yinmei Yang, Xingmei Wang, Qingyuan Li, Xufei Luo, Toshio Fukuoka, Hyeong Sik Ahn, Myeong Soo Lee, Zhengxiu Luo, Enmei Liu, Yao-long Chen, on behalf of COVID-19 evidence and recommendations working group

## Abstract

**Background:** The aim of this review was to evaluate the efficacy and safety of antibiotic agents in children with COVID-19, as well as to introduce the present situation of antibiotics use and bacterial coinfections in COVID-19 patients.

**Methods:** We searched Cochrane library, Medline, Embase, Web of Science, CBM, Wanfang Data and CNKI from their inception to March 31, 2020. In addition, we searched related studies on COVID-19 published before March 31, 2020 through Google Scholar. We evaluated the risk of bias of included studies, and synthesized the results using a qualitative synthesis.

**Results:** Six studies met our inclusion criteria. Five studies on SARS showed an overall risk of death of 7.2% to 20.0%. One study of SARS patients who used macrolides, quinolones or beta lactamases showed that the mean duration of hospital stay was 14.2, 13.8 and 16.2 days, respectively, and their average duration of fever was 14.3, 14.0 and 16.2 days, respectively. One cohort study on MERS indicated that macrolide therapy was not associated with a significant reduction in 90-day mortality (adjusted odds ratio [OR] 0.84, 95% confidence interval [CI] 0.47-1.51, *P* = 0.56) and improvement in MERS-CoV RNA clearance (adjusted hazard ratio [HR] 0.88, 95% CI 0.47, -1.64], *P* = 0.68). According to the findings of 33 studies, the proportion of antibiotics use ranged from 19.4% to 100.0% in children and 13.2% to 100.0% in adults, despite the lack of etiological evidence. The most commonly used antibiotics in adults were quinolones, cephalosporins and macrolides and in children meropenem and linezolid.

**Conclusions:** The benefits of antibiotic agents for adults with SARS or MERS were questionable in the absence of bacterial coinfections. There is no evidence to support the use of antibiotic agents for children with COVID-19 in the absence of bacterial coinfection.

## Background

In December 2019, an unexplained pneumonia emerged in Wuhan, China, and has since then spread rapidly throughout the country and the world. On February 11, the International Committee on Taxonomy of Viruses (ICTV) named the virus Severe Acute Respiratory Syndrome Coronavirus 2 (SARS-CoV-2) (1). On the same day, the World Health Organization (WHO) named the disease caused by the virus as COVID-19 (2). According to the latest data, by March 31 2020, a total of 1,174,866 cases had been confirmed worldwide, with 36,405 deaths. The number of confirmed cases in children continues to increase, with the youngest infected person being diagnosed only several minutes after birth (3). The outbreak of COVID-19 is the third introduction of a highly pathogenic coronavirus into the human population in the twenty-first century, after the Severe Acute Respiratory Syndrome (SARS) and Middle East Respiratory Syndrome (MERS) epidemics (4).

At present, there are no standardized or specific treatment schemes for COVID-19 patients, and the clinical treatment mainly focuses on symptomatic and supportive care. In principle, antibiotic agents should not be applied for viral infectious diseases like COVID-19, unless there is a bacterial coinfection. Besides, children have a higher risk of antibiotic-related adverse events than adults because of their anatomical and physiological features, particularly in the first years of life (5). Timely use of antibiotics is however needed when secondary bacterial infections are confirmed or community-acquired infections cannot be excluded (6-11). The potential overuse of antibiotics in patients infected with SARS-CoV-2 has become a major concern. Seven guidelines for COVID-19 have consistently pointed out that the unnecessary use of antibiotic agents, especially combinations of broad-spectrum antibiotics, should be avoided. According to a case series of 1099 patients with COVID-19, a majority of the patients (58.0%) received intravenous antibiotic therapy (12). Moreover, multiple case series and a cohort study of SARS patients testified that the proportion of patients receiving antibiotic agents used in the absence of a confirmed bacterial coinfection was between 50.0% and 100.0%, among whom 50.0% to 96.0% were treated with a combination of several antibiotic agents. The use of antibiotics over a long duration and in combinations of multiple agents not only showed no efficacy to the disease progress, but also caused complications such as potentially fatal secondary infections (13-18). However, some authors have argued that prophylactic use of antibiotics in the early stage can play a role in preventing infections in SARS patients (19). One study on SARS patients reported one patient recovered after receiving antibacterial treatment alone, and the condition of other patients improved after comprehensive treatment including antibiotics (20).

It is undisputed that antibiotic agents are essential for the treatment of confirmed bacterial infections, but whether antibiotic agents should be used to treat children with COVID-19 still remains a controversial issue. Therefore, we performed a rapid review to assess the value of antibiotic agents in children with COVID-19 and provide supporting evidence for the Rapid Advice Guideline for Management of Children with COVID-19. In addition, we intended to evaluate the current condition of the use of antibiotics and secondary infections in patients with COVID-19.

## Methods

### Search strategy

About the value of antibiotic agents in children with COVID-19, the following seven electronic databases were searched: Cochrane library, MEDLINE (via PubMed), Embase, Web of Science, CBM (China Biology Medicine disc), CNKI (China National Knowledge Infrastructure), and Wanfang Data up to March 31, 2020. The main terms were “2019-novel coronavirus”, “COVID-19”, “Middle East Respiratory Syndrome”, “Severe Acute Respiratory Syndrome” and “antibiotic agents” and so on (*The details of the search strategy can be found in the Supplementary Material 1*). We also searched clinical trial registry platforms (the World Health Organization Clinical Trials Registry Platform (http://www.who.int/ictrp/en/), US National Institutes of Health Trials Register (https://clinicaltrials.gov/), ISRCTN Register (https://www.isrctn.com/)), Google Scholar (https://scholar.google.nl/), and reference lists of all included publications for further potential studies.

About the current condition of the use of antibiotics and bacterial coinfections in patients with COVID-19, we searched all case series, case reports and descriptive studies related to COVID-19 published before March 31, 2020 through Google Scholar.

### Inclusion and exclusion criteria

To assess the current condition of the use of antibiotics and bacterial coinfections in patients with COVID-19, we included all case series, case reports and descriptive studies related to COVID-19, which reported the information on antibiotics use or bacterial coinfections. However, we ruled out studies without clear description on whether antibiotics were used for treating COVID-19, such as routinely use antibiotics after surgery or antibiotics used as eye drops. To assess the value of antibiotic use in children with COVID-19, we included studies that met the following criteria: 1) Types of studies: We primarily considered all types of studies about the use of antibiotics to treat patients with COVID-19. If we failed to identify sufficiently many studies, we also included studies about using antibiotics to treat SARS and MERS. 2) Types of participants: Studies including patients diagnosed with COVID-19 (and SARS and MERS if necessary), without restrictions on age, race, gender, geographical location or setting, were included. 3) Types of interventions: We included studies that compared the outcomes between patients taking antibiotic agents and those not. The types of antibiotics were not limited. We also included case series and case reports on comprehensive treatment with antibiotics and other drugs. Studies that only mentioned antibiotic treatment without explaining the specific methods of use and treatment effects were excluded. 4) Types of outcomes: The primary outcomes were mortality, duration of hospitalization and duration of fever. Secondary outcomes included chest X-ray absorptivity and other relevant indicators mentioned in the included studies.

We excluded: 1) animal studies and in vitro experiments; 2) studies not published in English or Chinese; 3) duplicates; or 4) conference abstracts, comments, and similar documents.

### Study selection

After eliminating duplicates, two reviewers performed independent searches in two steps as described below. Discrepancies were settled by discussion or consultation with a third reviewer. We used the bibliographic software EndNote. Prior to the formal selection, a training exercise of a random sample of 50 citations was conducted to ensure the reliability and feasibility of selection, until sufficient agreement on the selecting methods was reached.

In Step 1, all titles and abstracts were screened using pre-defined criteria. Studies were categorized into three groups (potentially eligible, excluded, and unclear). In Step 2, full-texts of potentially eligible and unclear studies were reviewed to identify the final inclusion. All reasons for exclusion of ineligible studies were recorded, and the process of study selection was documented using a PRISMA flow diagram (21).

### Data extraction

Two reviewers extracted the data independently with a standard data collection form. Any disagreements were resolved by consensus, and a third reviewer checked the consistency and accuracy of the data. Before the formal extraction, the form was piloted on a random sample of three included studies. The extraction form was finalized after counselling with clinicians.

Data extracted included: 1) Basic information: title, first author, publication year, study design and sample size; 2) Participants: baseline characteristics and disease of patients; 3) Details of the intervention and control conditions; and 4) Outcomes. For dichotomous outcomes, we abstracted the number of events and total number participants in each group; for continuous outcomes, we abstracted means, standard deviations (SD), and the number of total participants in each group. Outcomes with zero events were reported, but excluded from analysis.

About the case series and case reports of the use of antibiotics and bacterial coinfections in patients with COVID-19, we extracted basic information and the types and ratio of antibiotics and bacterial coinfections.

### Risk of bias assessment

We applied the Cochrane risk-of-bias (RoB) tool (22) for RCTs, Newcastle-Ottawa Scale(NOS) (23,24) for cohort studies and case-control studies, and the criteria recommended by the National Institute of Health and Clinical Optimization (NICE) for case series to assess the risk of bias (25). Two reviewers assessed the risk of bias independently following the overall assessment principle and disagreements were discussed in a consensus meeting. We used the above tools to produce a “Risk of Bias” summary table that included items, judgements, and support for judgements.

We did not assess the quality of case series and case reports on the use of antibiotics and bacterial coinfections in patients with COVID-19.

### Data synthesis

If the data were similar enough to be summarized in a meaningful way, we would conduct a meta-analysis using Review Manager 5.3. We would use fixed-effect meta-analysis for combining data where it was reasonable to assume that studies were evaluating the same underlying treatment effect, that was where trials all took the same intervention, and the trials’ populations and methods were judged to be sufficiently similar. If the clinical heterogeneity was sufficient enough to expect that the underlying treatment effects differ between trials, or if statistical heterogeneity was detected (*I*^2^ statistic > 50%), we used a random-effects meta-analysis to produce an overall summary, on the condition that an average treatment effect across trials was considered clinically meaningful. A qualitative synthesis was performed when significant heterogeneity existed.

For the case series and case reports on the use of antibiotics and bacterial coinfections in patients with COVID-19, we only described the current situation.

### Quality of the evidence assessment

Two reviewers (Jianjian Wang and Yuyi Tang) assessed the quality of evidence independently by using the Grading of Recommendations Assessment, Development and Evaluation (GRADE) approach. In the GRADE approach, direct evidence from RCTs begins at high quality, and evidence from observational studies at low level. The quality can be downgraded for five reasons (study limitations, consistency of effect, imprecision, indirectness, and publication bias) and upgraded for three reasons (large magnitude of effect, dose-response relation, and plausible confounders or biases) (26-31). In order to reflect the extent of our confidence that the estimates of the effect are correct, the quality of evidence will be graded as high, moderate, low, or very low. We produced a “Summary of Findings” table, which presented the overall quality of a body of evidence for each outcome, by using the GRADEpro software (32,33). We did not assess the quality of evidence of the case series and case reports on the use of antibiotics and bacterial coinfections in patients with COVID-19.

As COVID-19 is a public health emergency of international concern and the situation is evolving rapidly, our study was not registered in order to speed up the process (34).

## Results

### Clinical effects and safety of antibiotics

#### Study results

The literature search (*Figure 1*) yielded 2183 relevant records of studies on the value of the use of antibiotic agents (including studies on SARS and MERS), 778 of which were duplicates. After application of the exclusion criteria, six studies were included in the qualitative synthesis. There were no studies on COVID-19, therefore, we included studies on SARS and MERS fulfilling the inclusion criteria.

**Figure 1.**
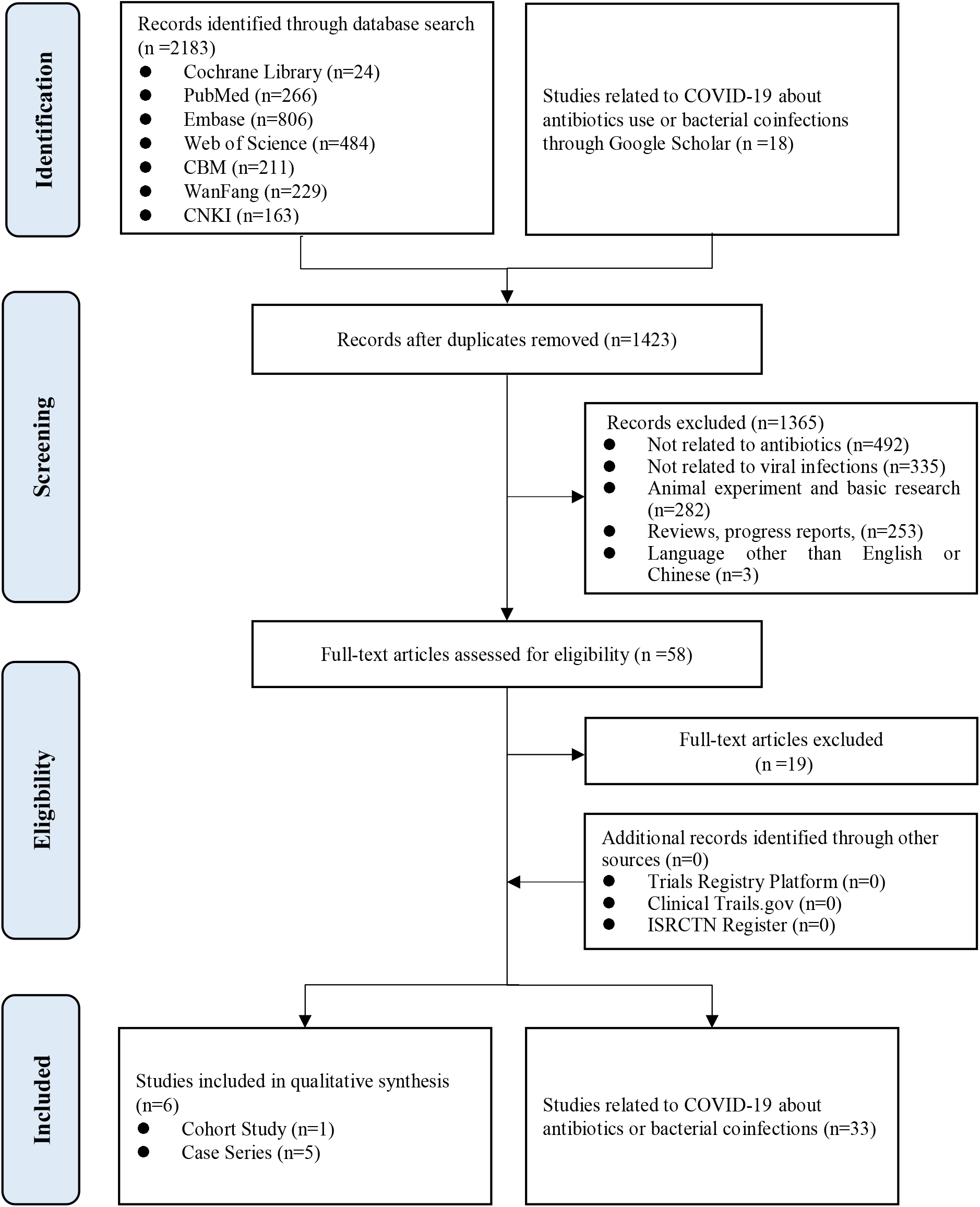
Flow diagram of the literature search.

#### Overview of studies

The six included studies were published between 2003 and 2019 (18,35-39). Sample sizes ranged between 10 and 349, with a total of 626 participants (*Table 1*). All participants of the six studies were adults; no studies on children were found. Five studies investigated SARS patients and one study enrolled patients with MERS. Due to the significant heterogeneity, we only conducted qualitative synthesis instead of a meta-analysis.

**Table 1.**
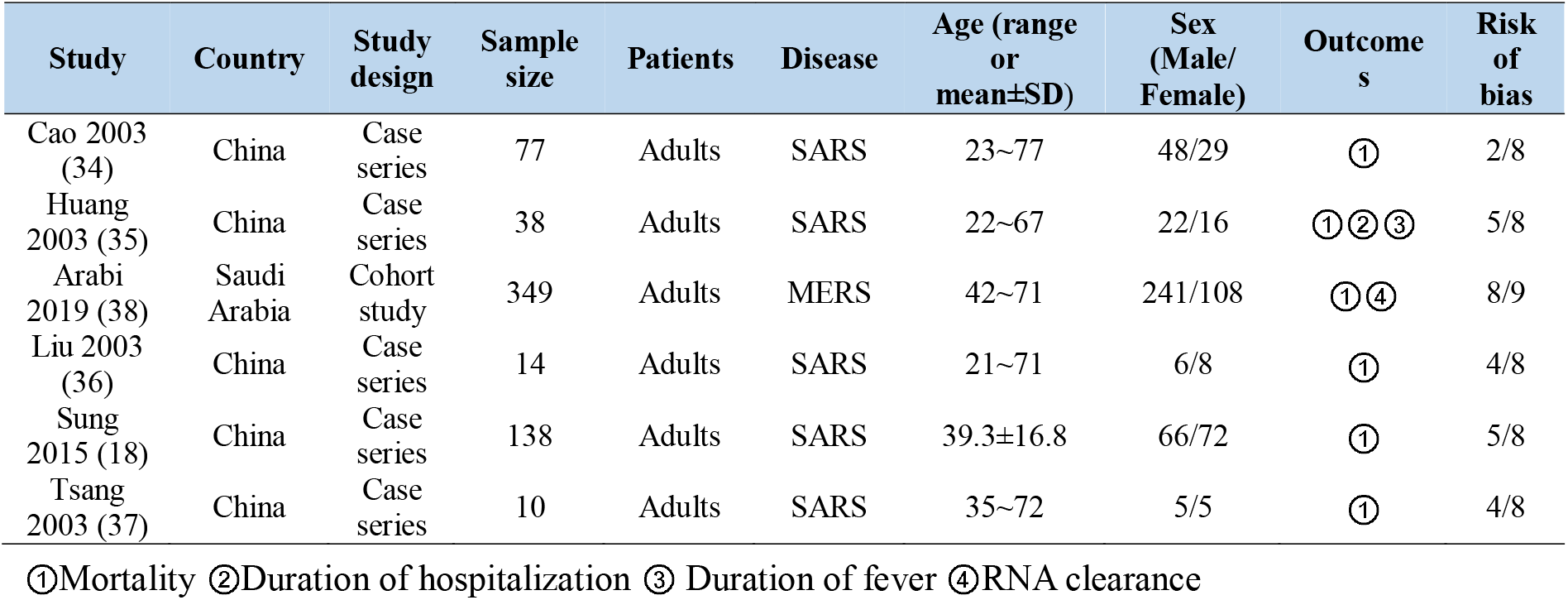
Characteristics and risk of bias of the included studies

#### Risk of bias

An overview of the quality of the included studies is presented in *Table 1*. Four case series had a moderate risk (score 4 to 5 out of 8), one case series had a high risk (score 2 out of 8), and the cohort study had a low risk (score 8 out of 9).

#### Summary of included studies Mortality

Mortality was the main outcome of all studies. In one study on SARS patients who took cephalosporin, macrolides and some broad-spectrum antibiotics, the risk of death was 13.0% (35). One case series of SARS patients showed that the risk of death in patients who used macrolides, quinolones and beta lactamases separately was 6.7%, 3.3% and 13.0%, respectively (36). Another case series of SARS patients showed a risk of death of 14.3% after using antibiotic agents for patients with confirmed secondary infection (37). One case series of patients using broad-spectrum antibiotics in the first two days, and another case series where the participants used a combination of beta-lactams and macrolide, showed overall risk of death of 10.9% and 20.0%, respectively (18,38). For MERS patients (39), there was no difference in the risk of death between patients treated with or without macrolide (adjusted OR=0.84, 95% CI [0.47, 1.51], *P*=0.56).

#### Duration of hospitalization

Only one study of SARS patients investigated the duration of hospital stay. In patients who used macrolides, quinolones or beta lactamases, the mean duration of hospital stay was 14.2, 13.8, and 16.2 days, respectively (36).

#### Duration of fever

One case series investigated the time from hospitalization until reaching normal body temperature in SARS patients. Patients treated with macrolides, quinolones or beta lactamases had an average fever duration of 14.3, 14.0, or 16.2 days, respectively (36).

#### RNA clearance

One study of MERS patients revealed that macrolide therapy was not associated with MERS-CoV RNA clearance (adjusted HR=0.88, 95% CI [0.47, 1.64], *P* = 0.68) (39).

#### Quality of evidence

For studies on SARS, the quality of evidence on mortality, duration of hospitalization and duration of fever was very low. The main reason was the risk of bias. For studies on MERS, the quality of evidence for the RNA clearance and 90-day mortality was low (*Table 2*).

**Table 2.**
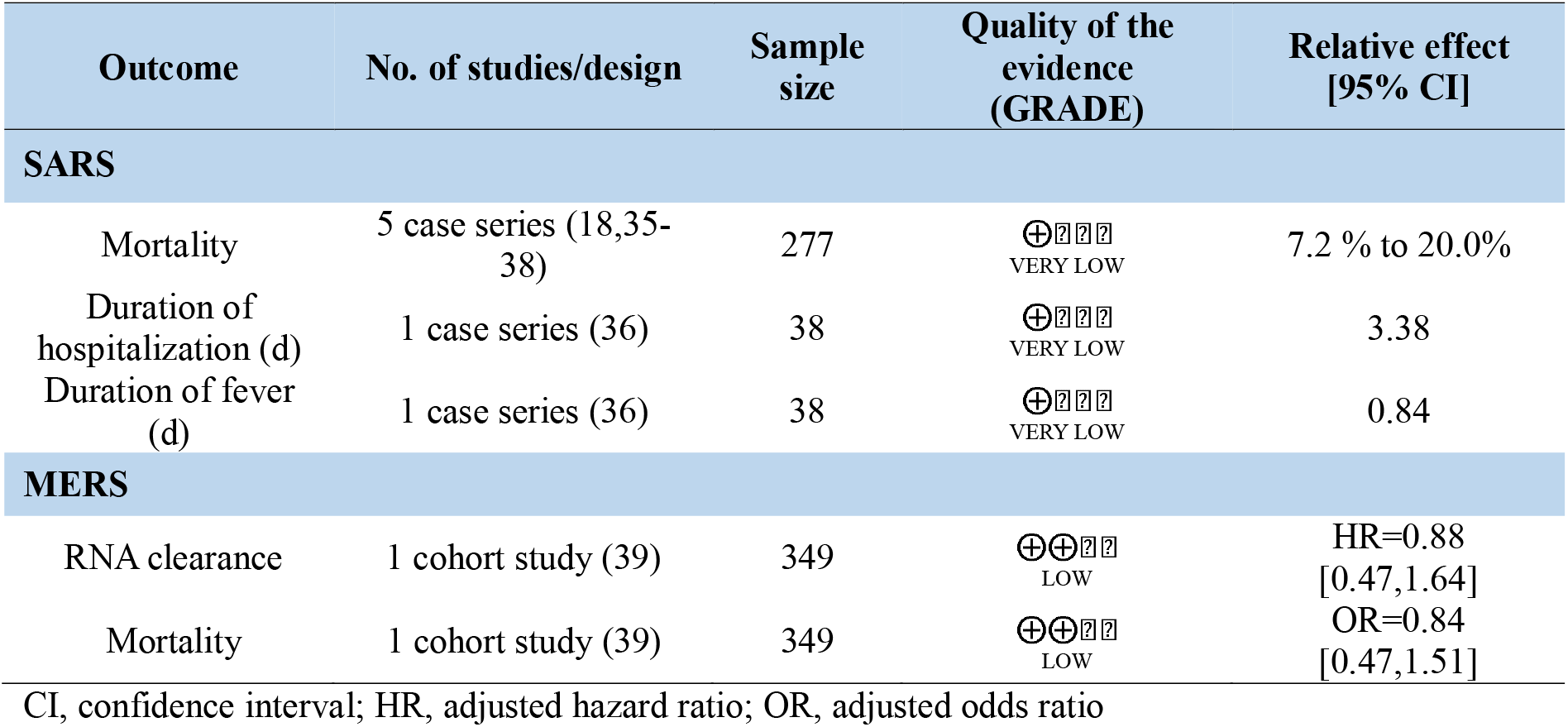
Quality of evidence

### Antibiotic agents and bacterial coinfection in studies on COVID-19

We identified 33 studies on COVID-19 that reported on antibiotic use or bacterial coinfections. Four studies on children with COVID-19 revealed that the proportion of the use of antibiotics ranged from 19.4% to 100.0% (*Table 3*). The most commonly used types were meropenem and linezolid. However, none of the four studies mentioned etiological findings. Twenty-nine studies of COVID-19 in adults showed that 13.2% to 100.0% of patients received antibiotics during hospital stay. The most commonly used types in adults were quinolones (especially moxifloxacin) cephalosporin and macrolides. However, only 1.0% to 27.3% of the patients had bacterial coinfections and the most common pathogens were Gram-negative bacillus such as *Acinetobacter baumannii, Klebsiella pneumoniae* and *Haemophilus influenzae*. (12,40-71).

**Table 3.** Antibiotic agents and bacterial coinfections in studies on COVID-19

| Study | Study design | Patients | Sample size | antibiotic agents |  | bacterial coinfection |  |
| --- | --- | --- | --- | --- | --- | --- | --- |
|  |  |  |  | Types | Number (percentage) | Types | Number (%) |
| Wang D, 2020 (40) | Case series | Children | 31 | / | 6 (19.4%) | / | / |
| Chen F, 2020 (41) | Case report | Children | 1 | Meropenem, Linezolid | 1 (100.0%) | / | / |
| Cai JH, 2020 (42) | Case report | Children | 10 | / | 5 (50.0%) | / | / |
| Liu WY, 2020 (43) | Case report | Children | 6 | / | 6 (100.0%) | / | / |
| Xu XW, 2020 (44) | Case series | Adults | 62 | Quinolones, Second-generation cephalosporin | 28 (45.2%) | / | / |
| Chen NS, 2020 (45) | Case series | Adults | 99 | Cefoperazone, Quinolones, Carbapenems, Tigecycline, Linezolid | 70 (70.7%) | Acinetobacter baumannii, Klebsiella pneumoniae | 1 (1.0%) |
| Chen L, 2020 (46) | Case series | Adults | 29 | / | / | / | 1 (3.4%) |
| Fang X, 2020 (47) | Case series | Adults | 79 | Moxifloxacin, Levofloxacin, Cefoperazone sulbactam | 49 (62.0%) | / | / |
| Wang Z, 2020 (48) | Case report | Adults | 4 | / | 4 (100.0%) | / | / |
| Wang D, 2020 (49) | Case series | Adults | 138 | Moxifloxacin, Ceftriaxone, Azithromycin | 138 (100.0%) | / | / |
| Guan W, 2020 (12) | Case series | Adults | 1099 | / | 637 (58.0%) | / | / |
| Wu 2020 (50) | Case series | Adults and children | 80 | Moxifloxacin | 73 (91.3%) | / | / |
| Liu 2020 (51) | Case series | Adults | 137 | / | 119 (86.9%) | / | / |
| Zhang 2020 (52) | Case report | Adults | 2 | Moxifloxacin | 2 (100.0%) | / | / |
| Huang 2020 (53) | Case series | Adults | 41 | / | 41 (100.0%) | / | 4 (10.0%) |
| Gao 2020 (54) | Case series | Adults | 11 | Moxifloxacin, Azithromycin | 3 (27.3%) | / | 3 (27.3%) |
| Zhao 2020 (55) | Case report | Adults | 2 | Moxifloxacin, Meropenem, Imipenem | 2 (100.0%) | / | / |
| Shi 2020 (56) | Case series | Adults | 67 | / | 22 (32.8%) | / | / |
| Easom 2020 (57) | Case series | Adults and children | 68 | Doxycycline, Moxifloxacin | 9 (13.2%) | Staphylococcus aureus, E.coli, Haemophilus influenzae | 4 (5.9%) |
| Chen 2020 (58) | Descriptive study | Adults | 274 | Moxifloxacin, Cefoperazone, Azithromycin | 249 (90.9%) | / | / |
| Chu 2020 (59) | Case series | Adults | 54 | / | 31 (57.4%) | / | / |
| Hayato 2020 (60) | Case report | Adults | 1 | Cefepime | 1 (100.0%) | / | / |
| Bai 2020 (61) | Case series | Adults | 58 | Levofloxacin, Moxifloxacin, Meropenem, Cefixime | 29 (50.0%) | / | / |
| Wang 2020 (62) | Case series | Adults | 67 | Moxifloxacin | 66 (98.5%) | Enterobacter cloacae, Acinetobacter baumannii | 5 (7.5%) |
| Chen 2020 (63) | Case series | Adults | 54 | Moxifloxacin, Cephalosporin, Carbapenems | 54 (100.0%) | / | / |
| Shang 2020 (64) | Case series | Adults | 36 | Moxifloxacin, Azithromycin, Piperacillin sulbactam | / | / | / |
| Cheng 2020 (65) | Cross-sectional study | Adults | 290 | Moxifloxacin, Levofloxacin, Cefdinir | 267 (92.1%) | / | / |
| Cheng 2020 (66) | Case series | Adults | 54 | Levofloxacin | 54 (100.0%) | / | / |
| Lei 2020 (67) | Case report | Adults | 9 | Cephalosporin, Azithromycin, Meropenem, Imipenem | 9 (100.0%) | / | / |
| Chen 2020 (68) | Case report | Adults and children | 9 | Moxifloxacin | 9 (100.0%) | / | / |
| Wan 2020 (69) | Case series | Adults | 135 | / | 59 (43.7%) | / | / |
| Ding 2020 (70) | Case report | Adults | 5 | / | 5 (100.0%) | / | / |
| Zhou 2020 (71) | Cohort study | Adults | 191 | / | 181 (95.0%) | / | 28 (15.0%) |

## Discussion

There is no direct evidence to support the efficacy of antibiotic agents in children with COVID-19. A high proportion of patients with COVID-19 were treated with antibiotics, despite the lack of etiological evidence. Current evidence suggests that secondary bacterial infections such as *Acinetobacter baumannii* and *Klebsiella pneumoniae* may occur in COVID-19 patients, and antibiotic agents are widely used in the clinical treatment of COVID-19. For SARS, evidence suggests that early use of antibiotics has no effect on the clinical outcomes, but the use of broad-spectrum antibiotics increased the risk of dysbacteriosis, which can cause nosocomial infections. However, when bacterial infections are identified, rational use of antibiotic agents showed valid results in relieving symptoms and reducing the leukocyte count.

The use of antibiotic agents is one of the most important clinical questions related to the management of COVID-19 in children. Most current guidelines suggest avoiding use of antibiotic agents blindly or inappropriately. They should only be used when there are confirmed secondary bacterial infections (72). Meanwhile, some guidelines and expert consensus statements believe that the use of antibiotic agents could be taken into consideration after glucocorticoid treatment, or if the patient has severe or critical illness, extensive lesion range, or large amount of airway secretions (73). A systematic review of non-severe pneumonia in children however found no evidence to support or question the continued use of antibiotic agents in children with non-severe pneumonia (74). Some researchers have emphasized the importance of toxic side effects and drug resistance to antibiotics (75), and many studies have also analyzed the bacterial types and drug resistance of secondary bacterial infections in SARS patients. Secondary infections are considered as an important risk factor for mortality in SARS patients (76,77).

This is the first systematic review aiming to assess the application of antibiotic agents for children with COVID-19. This study summarizes the literature on the efficacy and safety of antibiotic agents in the treatment of patients with COVID-19, SARS, and MERS. Both the amount and quality of the literature covered by this systematic review are however limited. This review mainly covered studies on the treatment of COVID-19 in adults and on SARS and MERS since we found no direct evidence for the treatment of COVID-19 in children. Due to the difference of patients’ inclusion criteria, treatment protocols, and outcomes measures among different studies, we were unable to carry out a meta-analysis. Therefore, high-quality clinical trials are needed to further confirm the efficacy of antibiotic agents for children with COVID-19. In addition, future studies should allow sufficient follow-up time and focus more on the adverse reactions to better evaluate the safety of antibiotic agents. Since the outbreak of COVID-19 is a public health emergency, in order to complete the rapid review as fast as possible, this review was not registered.

## Conclusions

In summary, we found no direct evidence to support the efficacy of antibiotic agents in children with COVID-19. Therefore, it remains unclear whether antibiotic agents should be used to treat children with COVID-19. Our rapid review showed that the benefits of antibiotics for adults infected with SARS or MERS was questionable in the absence of bacterial coinfections. We therefore recommend against the use of antibiotic agents for children with COVID-19 when there is no evidence of bacterial coinfection.

## Data Availability

This study is a rapid review based on the original researches. All the data are from the existing studies and are true.

## Author contributions

(I) Conception and design: Yaolong Chen and Enmei Liu; (II) Administrative support: Yaolong Chen; (III) Collection and assembly of data: Jianjian Wang and Yuyi Tang; (IV) Data analysis and interpretation: Jianjian Wang, Yuyi Tang and Yanfang Ma; (V) Manuscript writing: All authors; (VI) Final approval of manuscript: All authors.

## Acknowledgments

We thank Janne Estill, Institute of Global Health of University of Geneva for providing guidance and comments for our review. We thank all the authors for their wonderful collaboration.

## Footnote

### Ethical Statement

The authors are accountable for all aspects of the work in ensuring that questions related to the accuracy or integrity of any part of the work are appropriately investigated and resolved.

## Supplementary Material 1 Search strategy

### PubMed

#1. “COVID-19” [Supplementary Concept]

#2. “Severe Acute Respiratory Syndrome Coronavirus 2” [Supplementary Concept]

#3. “Middle East Respiratory Syndrome Coronavirus” [Mesh]

#4. “Severe Acute Respiratory Syndrome” [Mesh]

#5. “SARS Virus” [Mesh]

#6. “COVID-19” [Title/Abstract]

#7. “SARS-COV-2” [Title/Abstract]

#8. “Novel coronavirus” [Title/Abstract]

#9. “2019-novel coronavirus” [Title/Abstract]

#10 “coronavirus disease-19” [Title/Abstract]

#11 “coronavirus disease 2019” [Title/Abstract]

#12. “COVID 19” [Title/Abstract]

#13. “Novel CoV” [Title/Abstract]

#14.“2019-nCoV” [Title/Abstract]

#15. “2019-CoV” [Title/Abstract]

#16. “Wuhan-Cov” [Title/Abstract]

#17. “Wuhan Coronavirus” [Title/Abstract]

#18. “Wuhan seafood market pneumonia virus” [Title/Abstract]

#19. “Middle East Respiratory Syndrome” [Title/Abstract]

#20. “MERS” [Title/Abstract]

#21. “MERS-CoV” [Title/Abstract]

#22. “Severe Acute Respiratory Syndrome” [Title/Abstract]

#23. “SARS” [Title/Abstract]

#24. “SARS-CoV” [Title/Abstract]

#25. “SARS-Related” [Title/Abstract]

#26. “SARS-Associated” [Title/Abstract]

#27. #1-#26/ OR

#28. “Anti-Bacterial Agents”[Mesh]

#29. “Anti Bacterial Agents” [Title/Abstract]

#30. “Antibacterial Agents” [Title/Abstract]

#31. “Anti-Bacterial Compounds” [Title/Abstract]

#32. “Anti Bacterial Compounds” [Title/Abstract]

#33. “Bacteriocidal Agents” [Title/Abstract]

#34. “Bacteriocides” [Title/Abstract]

#35. “Anti-Mycobacterial Agents” [Title/Abstract]

#36. “Anti Mycobacterial Agents” [Title/Abstract]

#37. “Antimycobacterial” [Title/Abstract]

#38. “Antibiotic” [Title/Abstract]

#39. “Antibiotics” [Title/Abstract]

#40. “Antimicrobial Agents” [Title/Abstract]

#41. “Anti-infective agents” [Title/Abstract]

#42. “Monobactams”[Mesh]

#43. “Monobactams” [Title/Abstract]

#44. “Monobactam Antibiotics” [Title/Abstract]

#45. “Monocyclic beta-Lactams” [Title/Abstract]

#46. “Monocyclic beta Lactams” [Title/Abstract]

#47. “beta-Lactams”[Mesh]

#48. “Beta-lactam antibiotic*” [Title/Abstract]

#49. “Penicillins”[Mesh]

#50. “Penicillins” [Title/Abstract]

#51. “Penicillin Antibiotic*” [Title/Abstract]

#52. “Amoxicillin”[Mesh]

#53. “Amoxicillin” [Title/Abstract]

#54. “Cephalosporins”[Mesh]

#55. “Cephalosporins” [Title/Abstract]

#56. “Cephalosporin Antibiotics” [Title/Abstract]

#57. “Cephalosporanic Acids” [Title/Abstract]

#58. “Macrolides”[Mesh]

#59. “Macrolides” [Title/Abstract]

#60. “Azithromycin”[Mesh]

#61. “Azithromycin” [Title/Abstract]

#62. “Fluoroquinolones”[Mesh]

#63. “Fluoroquinolone” [Title/Abstract]

#64. “Tetracyclines”[Mesh]

#65. “Tetracyclines” [Title/Abstract]

#66. “Vancomycin”[Mesh]

#67. “Vancomycin” [Title/Abstract]

#68. #28-#67/ OR

#69. #27 AND #68

### Embase

#1. ‘ middle east respiratory syndrome coronavirus’/exp

#2. ‘severe acute respiratory syndrome’/exp

#3. ‘sars coronavirus’/exp

#4. ‘COVID-19’:ab,ti

#5. ‘SARS-COV-2’:ab,t0069

#6. ‘novel coronavirus’:ab,ti

#7. ‘2019-novel coronavirus’:ab,ti

#8. ‘coronavirus disease-19’:ab,ti

#9. ‘coronavirus disease 2019’:ab,ti

#10.‘COVID 19’:ab,ti

#11.‘novel cov’:ab,ti

#12.‘2019-ncov’:ab,ti

#13.‘2019-cov’:ab,ti

#14.‘wuhan-cov’:ab,ti

#15.‘wuhan coronavirus’:ab,ti

#16.‘wuhan seafood market pneumonia virus’:ab,ti

#17.‘middle east respiratory syndrome’:ab,ti

#18.‘middle east respiratory syndrome coronavirus’:ab,ti

#19.‘mers’:ab,ti

#20.‘mers-cov’:ab,ti

#21.‘severe acute respiratory syndrome’:ab,ti

#22.‘sars’:ab,ti

#23.‘sars-cov’:ab,ti

#24.‘sars-related’:ab,ti

#25.‘sars-associated’:ab,ti

#26.#1-#25/ OR

#27.‘antiinfective agent’/exp

#28.‘antibiotic agent’/exp

#29.‘anti bacterial agents’:ab,ti

#30.‘antibacterial agents’:ab,ti

#31.‘anti-bacterial compounds’:ab,ti

#32.‘anti bacterial compounds’:ab,ti

#33.‘bacteriocidal agents’:ab,ti

#34.‘bacteriocides’:ab,ti

#35.‘anti-mycobacterial agents’:ab,ti

#36.‘anti mycobacterial agents’:ab,ti

#37.‘antimycobacterial agent’/exp

#38.‘antimycobacterial’:ab,ti

#39.‘antibiotic’:ab,ti

#40.‘antibiotics’:ab,ti

#41.‘antimicrobial agents’:ab,ti

#42.‘anti-infective agents’:ab,ti

#43.‘monobactam derivative’/exp

#44.‘monobactams’:ab,ti

#45.‘monobactam antibiotics’:ab,ti

#46.‘monocyclic beta-lactams’:ab,ti

#47.‘monocyclic beta lactams’:ab,ti

#48.‘beta lactam antibiotic’/exp

#49.‘beta-lactam antibiotic*’:ab,ti

#50.‘penicillins’:ab,ti

#51.‘penicillin derivative’/exp

#52.‘penicillin antibiotic*’:ab,ti

#53.‘amoxicillin’/exp

#54.‘amoxicillin’:ab,ti

#55.‘cephalosporanic acid derivative’/exp

#56.‘cephalosporin’/exp

#57.‘cephalosporins’:ab,ti

#58.‘cephalosporin derivative’/exp

#59.‘cephalosporin antibiotics’:ab,ti

#60.‘cephalosporanic acids’:ab,ti

#61.‘macrolide’/exp

#62.‘macrolides’:ab,ti

#63.‘azithromycin’/exp

#64.‘azithromycin’:ab,ti

#65.‘quinolone derivative’/exp

#66.‘fluoroquinolone’:ab,ti

#67.‘tetracycline derivative‘’/exp

#68.‘tetracyclines’:ab,ti

#69.‘vancomycin’/exp

#70.‘vancomycin’:ab,ti

#71.#27-#70/ OR

#72.#26 AND #71

### Cochrane library

#1. MeSH descriptor: [Middle East Respiratory Syndrome Coronavirus] explode all trees

#2. MeSH descriptor: [Severe Acute Respiratory Syndrome] explode all trees

#3. MeSH descriptor: [SARS Virus] explode all trees

#4. “COVID-19”:ti,ab,kw

#5. “SARS-COV-2”:ti,ab,kw

#6. “Novel coronavirus”:ti,ab,kw

#7. “2019-novel coronavirus” :ti,ab,kw

#8. “Novel CoV” :ti,ab,kw

#9. “2019-nCoV” :ti,ab,kw

#10.“2019-CoV” :ti,ab,kw

#11.“coronavirus disease-19” :ti,ab,kw

#12.“coronavirus disease 2019” :ti,ab,kw

#13.“COVID 19” :ti,ab,kw

#14.“Wuhan-Cov” :ti,ab,kw

#15.“Wuhan Coronavirus” :ti,ab,kw

#16.“Wuhan seafood market pneumonia virus” :ti,ab,kw

#17.“Middle East Respiratory Syndrome” :ti,ab,kw

#18.“MERS”:ti,ab,kw

#19.“MERS-CoV”:ti,ab,kw

#20.“Severe Acute Respiratory Syndrome”:ti,ab,kw

#21.“SARS” :ti,ab,kw

#22.“SARS-CoV” :ti,ab,kw

#23.“SARS-Related”:ti,ab,kw

#24.“SARS-Associated”:ti,ab,kw

#25.#1-#24/ OR

#26.MeSH descriptor: [Anti-Bacterial Agents] explode all trees

#27.“Anti Bacterial Agents”:ti,ab,kw

#28.“Antibacterial Agents”:ti,ab,kw

#29.“Anti-Bacterial Compounds”:ti,ab,kw

#30.“Anti Bacterial Compounds”:ti,ab,kw

#31.“Bacteriocidal Agents”:ti,ab,kw

#32.“Bacteriocides”:ti,ab,kw

#33.“Anti-Mycobacterial Agents”:ti,ab,kw

#34.“Anti Mycobacterial Agents”:ti,ab,kw

#35.“Antimycobacterial”:ti,ab,kw

#36.“Antimycobacterial”:ti,ab,kw

#37.“Antibiotics”:ti,ab,kw

#38.“Antimicrobial Agents”:ti,ab,kw

#39.“Anti-infective agents”:ti,ab,kw

#40.MeSH descriptor: [Monobactams] explode all trees

#41.“Monobactams”:ti,ab,kw

#42.“Monobactam Antibiotics”:ti,ab,kw

#43.“Monocyclic beta-Lactams”:ti,ab,kw

#44.“Monocyclic beta Lactams”:ti,ab,kw

#45.MeSH descriptor: [beta-Lactams] explode all trees

#46.“Beta-lactam antibiotic”:ti,ab,kw

#47.MeSH descriptor: [Penicillins] explode all trees

#48.“Penicillins”:ti,ab,kw

#49.“Penicillin Antibiotic*”:ti,ab,kw

#50.MeSH descriptor: [Amoxicillin] explode all trees

#51.“Amoxicillin”:ti,ab,kw

#52.MeSH descriptor: [Cephalosporins] explode all trees

#53.“Cephalosporins”:ti,ab,kw

#54.“Cephalosporin Antibiotics”:ti,ab,kw

#55.“Cephalosporanic Acids”:ti,ab,kw

#56.MeSH descriptor: [Macrolides] explode all trees

#57.“Macrolides”:ti,ab,kw

#58.MeSH descriptor: [Azithromycin] explode all trees

#59.“Azithromycin”:ti,ab,kw

#60.MeSH descriptor: [Fluoroquinolones] explode all trees

#61.“Fluoroquinolone”:ti,ab,kw

#62.MeSH descriptor: [Tetracyclines] explode all trees

#63.“Tetracyclines”:ti,ab,kw

#64.MeSH descriptor: [Vancomycin] explode all trees

#65.“Vancomycin”:ti,ab,kw

#66.#26-#65/ OR

#67.#25 AND #66

## Web of Science

#1. TOPIC: “COVID-19”

#2. TOPIC: “SARS-COV-2”

#3. TOPIC: “Novel coronavirus”

#4. TOPIC: “2019-novel coronavirus”

#5. TOPIC: “coronavirus disease-19”

#6. TOPIC: “coronavirus disease 2019”

#7. TOPIC: “COVID 19”

#8. TOPIC: “Novel CoV”

#9. TOPIC: “2019-nCoV”

#10.TOPIC: “2019-CoV”

#11.TOPIC: “Wuhan-Cov”

#12.TOPIC: “Wuhan Coronavirus”

#13.TOPIC: “Wuhan seafood market pneumonia virus”

#14.TOPIC: “Middle East Respiratory Syndrome”

#15.TOPIC: “MERS”

#16.TOPIC: “MERS-CoV”

#17.TOPIC: “Severe Acute Respiratory Syndrome”

#18.TOPIC: “SARS”

#19.TOPIC: “SARS-CoV”

#20.TOPIC: “SARS-Related”

#21.TOPIC: “SARS-Associated”

#22.#1-#21/ OR

#23.TOPIC: “Anti-Bacterial Agents”

#24.TOPIC: “Anti Bacterial Agents”

#25.TOPIC: “Antibacterial Agents”

#26.TOPIC: “Anti-Bacterial Compounds”

#27.TOPIC: “Anti Bacterial Compounds”

#28.TOPIC: “Bacteriocidal Agents”

#29.TOPIC: “Bacteriocides”

#30.TOPIC: “Anti-Mycobacterial Agents”

#31.TOPIC: “Anti Mycobacterial Agents”

#32.TOPIC: “Antimycobacterial”

#33.TOPIC: “Antibiotic”

#34.TOPIC: “Antibiotics”

#35.TOPIC: “Antimicrobial Agents”

#36.TOPIC: “Anti-infective agents”

#37.TOPIC: “Monobactams”

#38.TOPIC: “Monobactam Antibiotics”

#39.TOPIC: “Monocyclic beta-Lactams”

#40.TOPIC: “Monocyclic beta Lactams”

#41.TOPIC: “Beta-lactam antibiotic*”

#42.TOPIC: “Penicillins”

#43.TOPIC: “Penicillin Antibiotic*”

#44.TOPIC: “Amoxicillin”

#45.TOPIC: “Cephalosporins”

#46.TOPIC: “Cephalosporin Antibiotics”

#47.TOPIC: “Cephalosporanic Acids”

#48.TOPIC: “Macrolides”

#49.TOPIC: “Azithromycin”

#50.TOPIC: “Fluoroquinolone”

#51.TOPIC: “Tetracyclines”

#52.TOPIC: “Vancomycin”

#53.#23-#52/ OR

#54.#22 AND #53

### CBM

#1. “新型冠状病毒”[常用字段:智能]

#2. “COVID-19”[常用字段:智能]

#3. “COVID 19”[常用字段智能]

#4. “2019-nCoV”[常用字段:智能]

#5. “2019-CoV”[常用字段:智能]

#6. “SARS-CoV-2”[常用字段:智能]

#7. “武汉冠状病毒”[常用字段:智能]

#8. “中东呼吸综合征冠状病毒”[不加权:扩展]

#9. “中东呼吸综合征”[常用字段:智能]

#10.“MERS”[常用字段:智能]

#11.“MERS-CoV”[常用字段:智能]

#12.“严重急性呼吸综合征&#x201D;[不加权:扩展]

#13.“SARS 病毒”[不加权:扩展]

#14.“严重急性呼吸综合征”[常用字段:智能]

#15.“SARS”[常用字段:智能]

#16.#1-#15/ OR

#17.“抗菌药”[不加权:扩展]

#18.“抗生素”[常用字段:智能]

#19.“抗菌药物”[常用字段:智能]

#20.“抗细菌药”[常用字段:智能]

#21.“抗菌素”[常用字段:智能]

#22.“内酰胺”[常用字段:智能]

#23.“ß-内酰胺酶”[常用字段:智能]

#24.“青霉素”[常用字段:智能]

#25.“青霉素类”[不加权:扩展]

#26.“阿莫西林”[常用字段:智能]

#27.“阿莫西林”[不加权:扩展]

#28.“头孢菌素类”[不加权:扩展]

#29.“头孢”[常用字段:智能]

#30.“大环内酯”[常用字段:智能]

#31.“大环内酯类”[不加权:扩展]

#32.“阿奇霉素”[常用字段:智能]

#33.“氟喹诺酮”[常用字段:智能]

#34.“四环素”[常用字段:智能]

#35.“四环素”[不加权:扩展]

#36.“万古霉素”[常用字段:智能]

#37.“万古霉素”[不加权:扩展]

#38.#17-#37/ OR

#39.#16 AND #38

### WanFang Data

#1. “新型冠状病毒”[主题]

#2. “COVID-19”[主题]

#3. “COVID 19”[主题]

#4. “2019-nCoV”[主题]

#5. “2019-CoV”[主题]

#6. “SARS-CoV-2”[主题]

#7. “武汉冠状病毒”[主题]

#8. “中东呼吸综合征”[主题]

#9. “MERS”[主题]

#10.“MERS-CoV”[主题]

#11.“严重急性呼吸综合征”[主题]

#12.“SARS”[主题]

#13.#1-#12/ OR

#14.“抗生素”[主题]

#15.“抗菌药物”[主题]

#16.“抗细菌药”[主题]

#17.“抗菌素”[主题]

#18.“内酰胺”[主题]

#19.“ß-内酰胺酶”[主题]

#20.“青霉素”[主题]

#21.“阿莫西林”[主题]

#22.“头孢”[主题]

#23.“大环内酯”[主题]

#24.“阿奇霉素”[主题]

#25.“氟喹诺酮”[主题]

#26.“四环素”[主题]

#27.“万古霉素”[主题]

#28.#14-#27/ OR #29.#13 AND #28

### CNKI

#1. “新型冠状病毒”[主题]

#2. “COVID-19”[主题]

#3. “COVID 19”[主题]

#4. “2019-nCoV”[主题]

#5. “2019-CoV”[主题]

#6. “SARS-CoV-2”[主题]

#7. “武汉冠状病毒”[主题]

#8. “中东呼吸综合征”[主题]

#9. “MERS”[主题]

#10.“MERS-CoV”[主题]

#11.“严重急性呼吸综合征”[主题]

#12.“SARS”[主题]

#13.#1-#12/ OR

#14.“抗生素”[主题]

#15.“抗菌药物”[主题]

#16.“抗细菌药”[主题]

#17.“抗菌素”[主题]

#18.“内酰胺”[主题]

#19.“ß-内酰胺酶”[主题]

#20.“青霉素”[主题]

#21.“阿莫西林”[主题]

#22.“头孢”[主题]

#23.“大环内酯”[主题]

#24.“阿奇霉素”[主题]

#25.“氟喹诺酮”[主题]

#26.“四环素”[主题]

#27.“万古霉素”[主题]

#28.#14-#27/ OR

#29.#13 AND #28

## References

1. Alexander EG, Susan CB, Ralph SB, et al. Severe acute respiratory syndrome-related coronavirus: The species and its viruses – a statement of the Coronavirus Study Group. BioRxiv 2020 [cited 2020 Apr 13]. Available online: https://www.biorxiv.org/content/10.1101/2020.02.07.937862v1

2. Novel Coronavirus (2019-nCoV) Situation Report-22[Internet]. World Health Organization; c2020 [cited 2020 Apr 13]. Available online: https://www.who.int/emergencies/diseases/novel-coronavirus-2019/situation-reports/

3. Novel Coronavirus (COVID-19) Situation[Internet]. World Health Organization; c2020 [cited 2020 Mar 31]. Available online: https://experience.arcgis.com/experience/685d0ace521648f8a5beeeee1b9125cd

4. de Wit E, van Doremalen N, Falzarano D, et al. SARS and MERS: recent insights into emerging coronaviruses. Nat Rev Microbiol 2016;14:523–34.

5. Principi N, Esposito S. Antibiotic-related adverse events in paediatrics: unique characteristics. Expert Opin Drug Saf 2019;18:795–802.

6. Clinical management of severe acute respiratory infection when novel coronavirus (nCoV) infection is suspected[Internet]. World Health Organization; c2020 [cited 2020 Apr 13]. Available online: https://www.who.int/docs/default-source/coronaviruse/clinical-management-of-novel-cov.pdf

7. Chen Z, Fu J, Shu Q, et al. Diagnosis and treatment recommendation for pediatric coronavirus disease-19. Journal of Zhejiang University (medical science) 2020;1–8.

8. The Society of Pediatrics of Hubei Medical Association. Recommendation for the diagnosis and treatment of novel coronavirus infection in children in Huber (Trail version 1). Chin J Contemp Pediatr 2020;22:96–9.

9. Jin Y, Cai L, Cheng Z, et al. A rapid advice guideline for the diagnosis and treatment of 2019 novel coronavirus (2019-nCoV) infected pneumonia (Standard version). Mil Med Res 2020;7:4.

10. Diagnosis and treatment of pneumonia caused by novel coronavirus (trial version 6)[Internet]. National Health Commission of People’s Republic of China; c2020 [cited 2020 Apr 13]. Available online: http://www.nhc.gov.cn/yzygj/s7653p/202002/8334a8326dd94d329df351d7da8aefc2/files/b218cfeb1bc54639af227f922bf6b817.pdf

11. Medical Expert Group of Tongji Hospital Affiliated to Tongji Medical College of Huazhong University of Science and Technology. A rapid advice guideline for the diagnosis and treatment of 2019 novel coronavirus infected pneumonia (Version 1). Herald Med 2020;1–9.

12. Guan W, Ni Z, Hu Y, et al. Clinical Characteristics of Coronavirus Disease 2019 in China. N Engl J Med 2020. [Epub ahead of print]

13. Li C, Pan S. Analysis and causation discussion of 185 severe acute respiratory syndrome dead cases. Chin Crit Care Med 2003;15:582–4.

14. Booth CM, Matukas LM, Tomlinson GA, et al. Clinical features and short-term outcomes of 144 patients with SARS in the greater Toronto area. JAMA 2003;289:2801–9.

15. Wei L, Duan D, Wang X, et al. Medical therapy and psychological treatment for SARS patients: experience with 51 cases. Chin J Pract Intern Med 2003;23:725–7.

16. Li W, Zheng Y, Huang K, et al. Complications at the anaphase of the critical severe acute respiratory syndrome and its treating strategies. Chin J Crit Care Med 2003;23:838–40.

17. Bleibtreu A, Jaureguiberry S, Houhou N, et al. Clinical management of respiratory syndrome in patients hospitalized for suspected Middle East respiratory syndrome coronavirus infection in the Paris area from 2013 to 2016. BMC Infect Dis 2018;18:331.

18. Sung JJ, Wu A, Joynt GM, et al. Severe acute respiratory syndrome: report of treatment and outcome after a major outbreak. Thorax 2004;59:414–20.

19. Wang P, Li M, Shi Y, et al. Evaluating the effects of different treatments on severe acute respiratory syndrome. Shanxi Med J 2005;34:270–2.

20. So LK, Lau AC, Yam LY, et al. Development of a standard treatment protocol for severe acute respiratory syndrome. Lancet 2003;361:1615–7.

21. Liberati A, Altman DG, Tetzlaff J, et al. The PRISMA statement for reporting systematic reviews and meta-analyses of studies that evaluate health care interventions: explanation and elaboration. J Clin Epidemiol 2009;62:e1–34.

22. Higgins JP, Altman DG, Gøtzsche PC, et al. The Cochrane Collaboration’s tool for assessing risk of bias in randomised trials. BMJ 2011;343:d5928.

23. Wells G, Shea B, O’Connell D, et al. New Castle-Ottawa Quality Assessment Scale — Cohort Studies. The Ottawa Hospital Research Institute, 2020 [cited 2020 Apr 13]. Available online: http://www.ohri.ca/programs/clinical_epidemiology/oxford.asp

24. Wells G, Shea B, O’Connell D, et al. New Castle–Ottawa Quality Assessment Scale — Case Control Studies. The Ottawa Hospital Research Institute, 2020 [cited 2020 Apr 13]. Available online: http://www.ohri.ca/programs/clinical_epidemiology/oxford.asp

25. Quality assessment for Case series[Internet]. National Institute for Health and Care Excellence; c2020 [cited 2020 Apr 13]. Available online: https://www.nice.org.uk/guidance/cg3/documents/appendix-4-quality-of-case-series-form2

26. Guyatt GH, Oxman AD, Vist G, et al. GRADE guidelines: 4. Rating the quality of evidence--study limitations (risk of bias). J Clin Epidemiol 2011;64:407–15.

27. Guyatt GH, Oxman AD, Montori V, et al. GRADE guidelines: 5. Rating the quality of evidence--publication bias. J Clin Epidemiol 2011;64:1277–82.

28. Guyatt GH, Oxman AD, Kunz R, et al. GRADE guidelines 6. Rating the quality of evidence--imprecision. J Clin Epidemiol 2011;64:1283–93.

29. Guyatt GH, Oxman AD, Kunz R, et al. GRADE guidelines: 7. Rating the quality of evidence--inconsistency. J Clin Epidemiol 2011;64:1294–1302.

30. Guyatt GH, Oxman AD, Kunz R, et al. GRADE guidelines: 8. Rating the quality of evidence--indirectness. J Clin Epidemiol 2011;64:1303–10.

31. Guyatt GH, Oxman AD, Sultan S, et al. GRADE guidelines: 9. Rating up the quality of evidence. J Clin Epidemiol 2011;64:1311–6.

32. Guyatt GH, Oxman AD, Akl EA, et al. GRADE guidelines: 1. Introduction-GRADE evidence profiles and summary of findings tables. J Clin Epidemiol 2011;64:383–94.

33. GRADEpro GDT. GRADEpro Guideline Development Tool[Software]. McMaster University, 2015 (developed by Evidence Prime, Inc.). Available online: www.gradepro.org

34. Ge L, Tian J, Li Y, et al. Association between prospective registration and overall reporting and methodological quality of systematic reviews: a meta-epidemiological study. J Clin Epidemiol 2018;93:45–55.

35. Cao B, Liu Z, Wang M, et al. Clinical diagnosis, treatment and prognosis of elderly patients with severe acute respiratory syndrome. Chin J Tuberc Respir Dis 2003;26:638.

36. Huang W, Xu H, Li Z, et al. 38 cases of severe acute respiratory syndrome. Chin J Infect Dis 2003;21:187–9.

37. Liu Z, Li T, Wang A, et al. The secondary infection in the treatment of patients with SARS. Chin Gen Prac 2003;552–3.

38. Tsang KW, Ho PL, Ooi GC, et al. A cluster of cases of severe acute respiratory syndrome in Hong Kong. N Engl J Med 2003;348:1977–85.

39. Arabi YM, Deeb AM, Al-Hameed F, et al. Macrolides in critically ill patients with Middle East Respiratory Syndrome. Int J Infect Dis 2019;81:184–90.

40. Wang D, Ju X, Xie F, et al. Clinical analysis of 31 cases of 2019 novel coronavirus infection in children from six provinces (autonomous region) of northern China. Chin J Pediatr 2020;58:1–7.

41. Chen F, Liu Z, Zhang F, et al. First case of severe childhood novel coronavirus pneumonia in China. Chin J Pediatr 2020;58:179–82.

42. Cai J, Xu J, Lin D, et al. A Case Series of children with 2019 novel coronavirus infection: clinical and epidemiological features. Clin Infect Dis 2020. [Epub ahead of print]

43. Liu W, Zhang Q, Chen J, et al. Detection of Covid-19 in Children in Early January 2020 in Wuhan, China. N Engl J Med 2020;382:1370–1.

44. Xu X, Wu X, Jiang X, et al. Clinical findings in a group of patients infected with the 2019 novel coronavirus (SARS-Cov-2) outside of Wuhan, China: retrospective case series. BMJ 2020;368:m606.

45. Chen N, Zhou M, Dong X, et al. Epidemiological and clinical characteristics of 99 cases of 2019 novel coronavirus pneumonia in Wuhan, China: a descriptive study. Lancet 2020;395:507–13.

46. Chen L, Liu H, Liu W, et al. Analysis of Clinical Features of 29 Patients With 2019 Novel Coronavirus Pneumonia. Chin J Tubere Respir Dis 2020;43:203–8.

47. Fang X, Mei Q, Yang T, et al. Clinical characteristics and treatment strategies of 79 patients with COVID-19. Chin Pharm Bull 2020;36:1–7.

48. Wang Z, Chen X, Lu Y, et al. Clinical characteristics and therapeutic procedure for four cases with 2019 novel coronavirus pneumonia receiving combined Chinese and Western medicine treatment. Biosci Trends 2020. [Epub ahead of print]

49. Wang D, Hu B, Hu C, et al. Clinical Characteristics of 138 Hospitalized Patients With 2019 Novel Coronavirus-Infected Pneumonia in Wuhan, China. JAMA 2020;e201585. [Epub ahead of print]

50. Wu J, Liu J, Zhao X, et al. Clinical Characteristics of Imported Cases of COVID-19 in Jiangsu Province: A Multicenter Descriptive Study. Clin Infect Dis 2020. [Epub ahead of print]

51. Liu K, Fang Y, Deng Y, et al. Clinical characteristics of novel coronavirus cases in tertiary hospitals in Hubei Province. Chin Med J, 2020. [Epub ahead of print]

52. Zhang Z, Li X, Zhang W, et al. Clinical Features and Treatment of 2019-nCov Pneumonia Patients in Wuhan: Report of A Couple Cases. Virol Sin 2020. [Epub ahead of print]

53. Huang C, Wang Y, Li X, et al. Clinical features of patients infected with 2019 novel coronavirus in Wuhan, China. Lancet 2020;395:497–506.

54. Gao T, He X, Su H, et al. Clinical characteristics of COVID-19: an analysis of 11 cases. Chin J Clin Infect Dis 2020;13:25–8.

55. Zhao X, Bao S, Du Y, et al. Diagnosis and treatment of 2 cases novel coronavirus pneumonia (2019 - nCoV) with acute respiratory failure. J Mod Oncol 2020;28:1415–8.

56. Shi J, Yang Z, Ye C, et al. Clinical observation on 49 cases of nonLJcritical coronavirus disease 2019 in Shanghai treated by integrated traditional Chinese and western medicine. Shanghai Journal of Traditional Chinese Medicine 2020;54:25–30.

57. Easom N, Moss P, Barlow G, et al. 68 Consecutive patients assessed for COVID-19 infection; experience from a UK regional infectious disease unit. Influenza Other Respir Viruses 2020. [Epub ahead of print]

58. Chen T, Wu D, Chen H, et al. Clinical characteristics of 113 deceased patients with coronavirus disease 2019: retrospective study. BMJ 2020;368:m1091.

59. Chu J, Yang N, Wei Y, et al. Clinical Characteristics of 54 medical staff with COVID-19: A retrospective study in a single center in Wuhan, China. J Med Virol 2020. [Epub ahead of print]

60. Hayato Taniguchi, Fumihiro Ogawa, Hiroshi Honzawa, et al. Veno - Venous Extracorporeal Membrane Oxygenation for Severe Pneumonia: COVID-19 Case in Japan. Wiley Online Library 2020 [cited 2020 Apr 13]. Available online: https://onlinelibrary.wiley.com/doi/10.1002/ams2.509

61. Bai P, He W, Zhang X, et al. Analysis of clinical features of 58 patients with severe or critical 2019 novel coronavirus pneumonia. Chin J Emerg Med 2020;29E022.

62. Wang Z, Yang B, Li Q, et al. Clinical Features of 69 Cases with Coronavirus Disease 2019 in Wuhan, China. Clin Infect Dis 2020. [Epub ahead of print]

63. Chen M, An W, Xia F, et al. Retrospective Analysis of COVID-19 Patients with Different Clinical Subtypes. Herald of Medicine 2020;1–12.

64. Shang X, Liu H, Zhu L, et al. Epidemiological and clinical characteristics of patients with coronavirus disease 2019 in FuYang city, AnHui province. Chinese Journal of Difficult and Complicated Cases 2020;1–5.

65. Cheng F, Li Q, Zeng F, et al. Analysis and recommendations of coronavirus disease 2019 related medication in Fangcang Hospital with 290 cases. Chinese Journal of Hospital Pharmacy 2020;1–4.

66. Cheng D, Li Y. Clinical Effectiveness and Case Analysis in 54 NCP Patients Treated with Lianhuaqingwen Granules. World Chinese Medicine 2020;1–5.

67. Lei D, Wang C, Li C, et al. Clinical characteristics of COVID-19 in pregnancy: analysis of nine cases. Chin J Perinat Med 2020;23:225–31.

68. Chen Q, Quan B, Li X, et al. A report of clinical diagnosis and treatment of 9 cases of coronavirus disease 2019. J Med Virol 2020. [Epub ahead of print]

69. Wan S, Xiang Y, Fang W, et al. Clinical Features and Treatment of COVID-19 Patients in Northeast Chongqing. J Med Virol 2020. [Epub ahead of print]

70. Ding Q, Lu P, Fan Y, et al. The clinical characteristics of pneumonia patients coinfected with 2019 novel coronavirus and influenza virus in Wuhan, China. J Med Virol 2020. [Epub ahead of print]

71. Zhou F, Yu T, Du R, et al. Clinical course and risk factors for mortality of adult inpatients with COVID-19 in Wuhan, China: a retrospective cohort study. Lancet 2020;395:1054–62.

72. Shen K, Yang Y, Wang T, et al. Diagnosis, treatment, and prevention of 2019 novel coronavirus infection in children: experts’ consensus statement. World J Pediatr 2020. [Epub ahead of print]

73. Xu K, Cai H, Shen Y, et al. Management of coronavirus disease-19 (COVID-19): the Zhejiang experience. Journal of Zhejiang University (medical science) 2020;1–12.

74. Lassi ZS, Kumar R, Das JK, et al. Antibiotic therapy versus no antibiotic therapy for children aged two to 59 months with WHO-defined non-severe pneumonia and wheeze. Cochrane Database Syst Rev 2014;CD009576.

75. Feng D. The research and analysis of future potential applications of the antibiotic. The Journal of Shandong Agricultural Engineering University 2016;33:35–40.

76. Li L, Shi Y, Liu H. Organism distribution and drug resistance analysis in 13 cases of SARS patients with secondary infection. Chin J Infect Control 2005;4:252–4.

77. Huang X, Zhuang J, Pang X, et al. Organism distribution and drug resistance in severe acute respiratory syndrome with secondary bacterial infection. Chin J Nosocomiol 2003;13:1084–6.

